# Development and validation of a tool to aid writing and reviewing of healthcare safety investigation reports: a modified-Delphi study

**DOI:** 10.1101/2025.11.26.25341045

**Authors:** Paul Bowie, Melanie Ottewill, Rosemary Lim, Tracey Herlihey, Ethel Oldfield, Helen Vosper, Ian Davidson, Hugh Currie, Jennifer O’Donnell, Nicola Steevenson, Helen Higham, Andrew Murphy-Pittock, Duncan McNab, Alexia Pellowe, Martin Duffy, Manoj Kumar, Tom McEwan, Elizabeth Crisp, Meredith Makeham, Catie Paton, Sarahjane Jones, Peter Hibbert, Andrew Carson-Stevens, Alastair Ross

## Abstract

**Introduction:** Healthcare organisations worldwide are expected to investigate or review incidents that unintentionally harm or could cause harm. Investigation findings are presented in a written report, which is arguably a proxy measure for the quality of investigations. The report is an important document to be read and acted upon. The aims of this study were: 1) to design a tool to support the writing and review of healthcare safety investigation reports, and 2) to validate the content of the tool.

**Methods:** The study was conducted in two phases. Phase 1: The initial content of the tool was developed using relevant published literature and building consensus with 23 specialist participants from the United Kingdom and Australia. Phase 2: Content validity of the tool was assessed for relevance and clarity in two e-Delphi survey rounds with users of the tool. Using a 4-point scale, a median of 3 or 4 and an interquartile range of less than or equal to 1.5 were used to determine consensus.

**Results:** Phase 1: A tool containing 8 areas of review with accompanying descriptors was developed. For each area of review, a 3-point ordinal rating scale along with a comments box for formative self-assessment were included. Phase 2: At the end of the round 1 survey, there was consensus on all but one area of review in the tool. By the end of round 2 survey, consensus was reached on all areas of review. No additional areas of review were added and none were removed. Some descriptors were amended to improve clarity.

**Conclusion:** We co-designed and validated the content of a tool that can be used to inform the quality of safety investigation reports and learning in terms of a systems-based approach. The tool has multiple uses ranging from self-assessment for report writers to facilitating oversight of the quality of healthcare safety investigation reports. Future work could focus on building further evidence of the tool’s overall utility.

**What is already known on this topic:** – summarise the state of scientific knowledge on this subject before you did your study and why this study needed to be done

- The standard of healthcare safety investigation and reports internationally is known to be variable and often lacks the use of a Human Factors informed “systems approach”.
- A formal mechanism appears to be lacking to facilitate a review of, and provide feedback on, the standard of healthcare investigation reports, which arguably serves as a proxy for the overall quality of the investigation process.

**What this study adds:** – summarise what we now know as a result of this study that we did not know before

- To our knowledge, this is a first validated tool that has been developed to support self-assessment and oversight of written safety investigations reports and learning reviews.
- Key principles of a systems-based approach to healthcare safety investigations and learning reviews are incorporated into a single tool to guide self-assessment and improvement, where needed.

**How this study might affect research, practice or policy:** – *summarise the implications of this study*

- The tool supports organisational quality assurance or oversight mechanisms for monitoring, evaluating and improving the standard of investigation reports.
- As written report can act as a proxy measure for the quality of investigations, improving the standard of reports may inform the learning and associated action from healthcare safety investigations.
- Further research and evaluation are necessary to provide greater evidence of the utility of the tool.

## Introduction

Healthcare organisations worldwide are routinely expected to investigate (or review), accidents and incidents that unintentionally harm, or could have harmed, patients and others [1–4]. Different approaches to investigation may be used [5–10]. But, whatever the approach, the aim is to identify learning for the purposes of improving patient safety [11–14]. The findings from the investigation will usually be presented in a written report, which details the event in question, the data collection approach taken, an analysis of the evidence gathered, identified learning for the organisation and recommendations for improvement or changes in practice [15]. The report is usually subject to scrutiny, review and ‘sign-off’ at senior leadership level. It is an important document and output of a specific investigation that will be read and possibly acted upon by a range of audiences including patients, families, carers, healthcare professionals, coroners and legal teams [1, 14]. The written report is arguably a proxy measure for the quality of the overall investigation process undertaken.

However, published evidence in healthcare highlights long-standing issues with the standard of healthcare safety investigations and the resulting written reports, with many limited in terms of taking a recognised “systems approach” as recommended by the Human Factors and Ergonomics profession [4, 6, 9, 16–18]. For example, recommendations for improvements are often considered ‘weak’ in terms of their capacity to reduce the risk of recurrence [19–20]. One barrier to the standard of both investigations and written reports is the availability and quality of training given to those leading and conducting safety investigations [21–22]. A further issue is the reported theoretical problems and assumptions underpinning the analytical framework approach to investigation that is often adopted, typically Root Cause Analysis [23–24]. Together with the weak risk control strategies resulting from these investigations [19–20], there also appears to be a lack of double-loop learning and feedback mechanisms that is ‘needed to secure change’ within organisations [25].

A formal mechanism to facilitate a formative review of, and provide feedback on, the standard of healthcare investigation reports from a systems perspective, and by proxy, the quality of the overall investigation process appears to be lacking. A similar approach, based on an external peer review process for significant event analyses [26–27], is established in primary care settings in the west of Scotland for medical educational purposes, but it is not widely applied beyond this region. There is a self-evident need to develop a supportive review and feedback process to address the well-recognised challenges with healthcare safety investigations and the resulting standard of written reports, demonstrable learning and improvements.

This type of tool could be used by writers of healthcare safety investigation reports, and those who review and sign off these reports. They can potentially use the tool to:

- assess the quality of the written report which, in turn, provides an indication of the quality of the investigation.
- highlight and share good practices
- identify areas for investigation process improvement
- inform training requirements to facilitate more meaningful investigations and reports

Against this background, the aims of this study were twofold:

1. To co-design a formative tool to support the writing and review of healthcare safety investigation reports.
2. To validate the content of the tool with expert and informed users.

## Methods

### Study Design

We used a two-phase modified-Delphi approach [28] by complemented by peer-reviewed and grey literature and content analysis of safety investigation reports [29]. Studies employing the Delphi technique recruit individuals who know about the topic being investigated, usually referred to as ‘experts’. There is a lack of agreement on the optimum number of participants to include on a Delphi panel, and it is recommended that sample size be dependent upon what is being investigated, the complexity of the problem, the homogeneity or heterogeneity of the sample, and the availability of resources.

As participants are relatively heterogenous, in line with sample size recommendations for a heterogenous sample i.e. up to a minimum of 10-18 participants, we aimed to recruit around 20 participants. This number is also to increase group judgement reliability and make up for anticipated participant attrition.

Typical response rates for online survey questionnaires are 10% [30] but we do not have any reliable estimates for response rates for our type of study. Given the niche topic area and lack of evidence base for the issue at hand, we anticipated that the study would be of high interest to those we approached to participate in this study. We took a flexible and pragmatic approach and did not exclude participants unnecessarily. Typically, a Delphi technique uses multiple iterations of data collection rounds, usually two or three, to gain agreement.

### Phase 1: Tool development

This phase was conducted over a 6-month period (October 2022 to March 2023) with participants representing a mix of regional and national healthcare organisations and higher education institutions in the UK and Australia.

### Study participants and recruitment

This phase was managed by three project leaders (PB, MO, TH), all of whom have significant experience (>10 years) in patient safety, investigation practice, education or research.

The project leads identified and recruited potential ‘experts’ via email to be part of the modified-Delphi panel. Up to 20 panel members were targeted for recruitment. ‘Expertise’ was accorded based on a combination of:

1. Being informed and experienced in safety investigation related practice, education or research.
2. Having published on patient safety learning issues in international peer reviewed journals;
3. Holding senior leadership roles in patient safety and/or related research or education; and
4. Having acquired >5 years’ experience of safety investigation practice.

### Tool content development

#### Preliminary development of the tool

The preliminary development of the tool was informed by:

- Study of the review of healthcare safety investigations and complaints reports by Human Factors specialists that led to the identification of ‘traps to avoid’ in safety investigation and learning reviews [29].
- A comprehensive search to identify key published literature that maybe helpful in informing ‘good practice’ principles in relation to healthcare safety investigation.
- A hand search of key healthcare journals (*BMJ Quality & Safety; BMJ Open Quality; Journal of Patient Safety; The Joint Commission Journal on Quality and Patient Safety; International Journal for Quality in Health Care; and the Journal of Patient Safety and Risk Management*)
- Professional cross sector ‘good practice’ guidance on learning from adverse events by the UK-based Chartered Institute of Ergonomics and Human Factors and the Energy Institute [16, 31].

After careful review and re-review of the identified literature, PB extracted key information from reported study results and discussion sections where it was judged investigation and learning review practices could be improved. From this, a preliminary draft tool was then created that comprised a series of 13 written statements that aligned to published evidence in support of their inclusion - alongside a simple ordinal rating scale to guide the formative self-assessment process.

### Iterative co-design of the tool

The preliminary draft tool was circulated to the expert panel via electronic mail for critical review and the provision of feedback. Panel members were asked to comment on the relevance of each statement for the formative review of safety investigation reports. Panel members were also asked to comment on the clarity of the statements and provide feedback for improvement. They were also asked to suggest additional items for the tool. The tool was iteratively redesigned based on three rounds of feedback received. PB made a judgement after each round to exclude items or edit them to enhance relevance and clarity in preparation for the next modified-Delphi round.

Disagreements were resolved by discussion via online meetings at the end of each round of feedback (using a combination of 1 to 1 meetings and small group meetings e.g. 3-6 participants at a time to accommodate availability) using Microsoft Teams until consensus was reached on the final content of the preliminary tool. At the end of this phase, the tool consisted of eight statements.

### Phase 2: Validating the content of the tool

*Study Design:* The study adopted a modified Delphi technique using questionnaires.

*Sample:*

Participants:

Inclusion criteria: current users of the tool and either

1. Healthcare safety investigators
2. Healthcare staff who have an oversight role for safety investigations

Exclusion criteria: Any individual who did not fit into any of the categories in the inclusion criteria.

### Sample size

Up to 20 participants.

### Recruitment and informed consent

Non-probability sampling methods (purposive and snowballing sampling techniques) were used to identify potential participants who fulfilled the inclusion criteria. Potential participants were identified by MO, PB and RL, who have key contacts in this area of research and practice (purposive sampling). Potential participants identified and/or recruited through purposive sampling were asked to suggest others whom they knew fulfilled the inclusion criteria (snowballing sampling). Only participants who gave informed consent were contacted further to complete the questionnaires.

### Data Collection

Participants completed two online questionnaires. The questionnaires asked the panel of participants to rate the eight statements in the tool (using a 4-point Content Validity Index) against two content validity criteria, which were ‘relevance’ and ‘clarity’ reduced from the four as described by Yaghmale [32] as these two were judged to be sufficient for the purposes of our study.

REDCap, a tool for creating web surveys was used to administer the questionnaires [https://project-redcap.org/]. Participants were sent an e-mail containing a link to the questionnaire. A reminder email was sent at two weeks to encourage timely submission of the completed questionnaire. The questionnaire was open for 4 weeks. After analysis of the responses, a weblink of the second round of the questionnaire was emailed to the same participants.

### Data Analysis

Survey data was analysed by calculating median scores and interquartile ranges (IQRs) for responses to each area of review in the tool. IQRs were used to assess the level of consensus for each question/area of review. A median score of 3 or 4 and an IQR range of ≤ 1.5 were considered as the cut off point for reaching consensus. Content analysis was used to explore qualitative responses obtained from the questionnaire.

## Results

### Phase 1 – Initial Tool Development

A total of 23 safety learning and human factors specialists, leaders, clinicians, educators and academic researchers, all with more than 10-years’ experience took part in the development of the tool (Table 1).

**Table 1.** Professional Characteristics of ‘Expert’ Development Group (n=23)

| **Expert No.** | **Professional Role(s)** | **Employing Organisation(s)** | **Experience in Patient Safety Learning (Yrs)** |
| --- | --- | --- | --- |
| 1.<br>(PB) | Human Factors Specialist/ Senior Patient Safety Researcher / Academic and Educator | NHS Education for Scotland / Health Services Safety Investigations Body, UK | 25+ |
| 2.<br>(MO) | Registered Nurse and Social Worker / Human Factors Specialist / Patient Safety Investigator | Health Services Safety Investigations Body, UK | 25+ |
| 3.<br>(TH) | Human Factors Specialist/ Senior Patient Safety Leader and Educator | NHS England Patient Safety Team | 15+ |
| 4.<br>(EO) | Midwife/Human Factors Specialist/Safety Investigation Specialist | NHS England Patient Safety Team, UK | 10+ |
| 5.<br>(HV) | Human Factors Specialist/ Senior Patient Safety Researcher / Academic and Educator | Health Services Safety Investigations Body / University of Aberdeen, UK | 15+ |
| 6.<br>(ID) | Senior Risk and Safety Leader | NHS Tayside Health Board, UK | 10+ |
| 7.<br>(HC) | Senior Risk and Safety Director / Occupational Safety and Health Specialist | NHS Ayrshire and Arran Health Board, UK | 20+ |
| 8.<br>(JOD) | Human Factors Specialist / Safety Investigation Specialist / Educator | Systemic Factors / City University, UK | 20+ |
| 9.<br>(NS) | Human Factors Specialist / Safety Investigation Specialist | Systemic Factors / University of Derby, UK | 10+ |
| 10.<br>(HH) | Medical Practitioner/ Patient Safety Research Director / Academic and Educator | University of Oxford, UK | 20+ |
| 11.<br>(AM-P) | Medical Educator/Director Safety Investigation Education | Health Services Safety Investigations Body, UK | 20+ |
| 12.<br>DMc | Medical Practitioner/ Senior Patient Safety Researcher / Academic and Educator | NHS Education for Scotland, UK | 20+ |
| 13.<br>(AP) | Medical Practitioner/ Senior Patient Safety Educator | NHS Education for Scotland, UK | 10+ |
| 14.<br>(MD) | Medical Practitioner/ Senior Patient Safety Educator | Belfast Health and Social Care Trust, UK | 15+ |
| 15.<br>(RL) | Pharmacist/Senior Patient Safety Researcher / Academic and Educator | Health Services Safety Investigations Body/ University of Reading, UK | 10+ |
| 16.<br>(MK) | Consultant Surgeon/National Patient Safety Leader | NHS Education for Scotland<br>NHS Grampian, UK | 15+ |
| 17.<br>(ACS) | Medical Practitioner/ Patient Safety Professor /<br>Academic and Educator | Cardiff University, UK | 15+ |
| 18.<br>(TMC) | Clinician/Safety Investigation Education/Academic | NHS Education for Scotland, UK | 10+ |
| 19.<br>(EC) | Senior Midwife, Academic Researcher, Safety<br>Scientist | Staffordshire University, UK | 10+ |
| 20,<br>(MM) | Medical Practitioner/ Patient Safety Professor /<br>Academic and Educator | Sydney University, Australia | 15+ |
| 21.<br>(SJJ) | Medical Scientist/ Patient Safety Professor /<br>Academic, Researcher and Educator | Staffordshire University, UK | 10+ |
| 22.<br>(PH) | Patient Safety Professor / Academic, Researcher<br>and Educator | Centre for Healthcare Resilience and<br>Implementation Science, Australia | 15+ |
| 23.<br>(AR) | Human Factors Specialist/ Senior Patient Safety<br>Researcher / Academic | Staffordshire University, UK | 20+ |

The development and consensus building process went through three iterations of the modified-Delphi process until final agreement was reached. During this process, the tool content was reduced from 13 items to 8 items. For example, “t*he need to consider the importance of both ‘context’ and ‘situation’*” in the tool was removed due to lack of sufficient agreement by the group. Similarly, “*the need to acknowledge the difference between intentional and unintentional human failure*”, and “*do not confuse recommendations with solutions*” were considered but eventually omitted due to limited agreement on their perceived usefulness. Consideration was also given to potential missing items (e.g. using safety-related terms in the wrong context). These items were suggested by panel members, but these were either rejected following initial discussions (online and via email) or the necessary level of agreement was not achieved because of the potential for over-complicating the tool.

Different types of rating scales were suggested but failed to garner consensus. A ‘written comments’ section was added for each item. The purpose of these sections was for users to:

- clarify and rationalise their ratings,
- highlight areas that could be improved
- highlight content that was thought to have worked well and should be shared with others for standard-setting future reports and to inform local training.
- Include direct quotes to explicitly highlight good practice or learning opportunities.

Multiple minor language edits took place to improve relevance, readability, simplicity and clarity of the tool content. At the end of the 3-round feedback, an 8-item tool with descriptors and a 3-point ordinal rating scale (“good evidence; “some evidence”, “little evidence”) (see Table 2) was agreed.

**Table 2.** Preliminary Content of the Learning Response Review Tool.

| **Item No** | **Item** | **Descriptor** | **Supporting Evidence** |
| --- | --- | --- | --- |
| **1** | **People affected by incidents are meaningfully engaged and involved** | The report demonstrates evidence that all those affected by the incident such as staff, patients, families and carers have been actively listened to and emotionally supported where required (i.e. interviews and perspectives of those affected are included in the report). | 21-22, 50-54 |
| **2** | **The “Systems Approach” is applied** | The report demonstrates consideration of system-based performance influencing factors (e.g. task complexity, technology, work procedures, workplace design, information transfer, clinical condition of patient, stress, fatigue, culture, leadership/management, policy/regulation) and how these interacted to contribute to the incident in question. | 16-17, 31, 33, 37-39, 55-57 |
| **3** | **‘Human Error’ is considered as a symptom of a system problem** | ‘Human error’ or similar (e.g. nurse error, medical error, loss of situation awareness) is not concluded to be the ‘cause’ of the incident. Instead, multiple contributory factors which influenced the event are acknowledged and explored. | 16, 43, 58-60 |
| **4** | **Blame language is avoided** | Language does NOT directly or indirectly infer blame of individuals, teams, departments, or organisations and/or focus on human failure (i.e. the nurse failed to follow policy; the doctor lost ‘situation awareness’). | 16, 59, 61-62 |
| **5** | **Local Rationality is considered** | The report clearly explains why the decisions and actions taken by individuals involved felt right at the time (i.e. the system situation and context faced by those individuals are explored and described). | 16, 29, 63 |
| **6** | **Counterfactual reasoning is avoided** | The report focuses on what happened and understanding why an incident happened. The report does NOT make a judgement on what people, departments or organisations ‘could’ or ‘should’ have done during or before the incident. | 16, 28, 63 |
| **7** | **Safety actions/recommendations are systems-based, developed collaboratively and evidence-based** | Safety actions/recommendations proposed:<br>have been developed collaboratively with relevant staff/stakeholders and with consideration of wider organisation priorities and improvement work.<br>focus on system elements (IT, equipment, care processes/pathways) not individuals<br>are specific, robust and actionable i.e. they don’t add to ‘safety clutter’<br>are accompanied by a plan to monitor progress over time<br>are demonstrably linked to the evidence and findings in the report | 16, 19-20, 23, 45-48, |
| 8 | **The written report is clear, easy to read and anonymised** | The report is concise, written in plain English, uses inclusive language and anonymised i.e. it is written to 'inform rather than impress' | 10, 16, 31, 33 |

The tool was named the ‘Learning Response Review and Improvement Tool’. The reasoning was to reflect the updated policy direction advocated by NHS England’s ‘Patient Safety Incident Response Framework (PSIRF), which aims to support a ‘systems based’ to investigating and learning from patient safety incidents [33]. In England, the term “Learning response” is used to encompass the different analytical and learning methods that can be applied such as Swarm Huddles, After Action Reviews and Patient Safety Incident Investigations [34].

### Phase 2: Content Validity

#### Demographics

A total of 17 participants completed the round 1 survey, and 11 completed the round 2 survey. Participants in round 1 worked in the UK (n=15) and Australia (n=2). They had been involved in healthcare safety investigations for an average of 7 years (range, 2 months to 20 years), with job roles such as director of patient safety, head of safety investigations, patient safety investigator, patient safety specialist and researchers. On average, participants in round 1 used the tool 17 times (range 1-85).

#### Round 1 Results

There was consensus on all areas of review in terms of relevance and clarity except for ‘clarity’ in area of review 1 (see Table 3). Participants also provided comments on each area of review. The research team considered each comment and made changes to the wordings to all 8 areas of review.

**Table 3:**
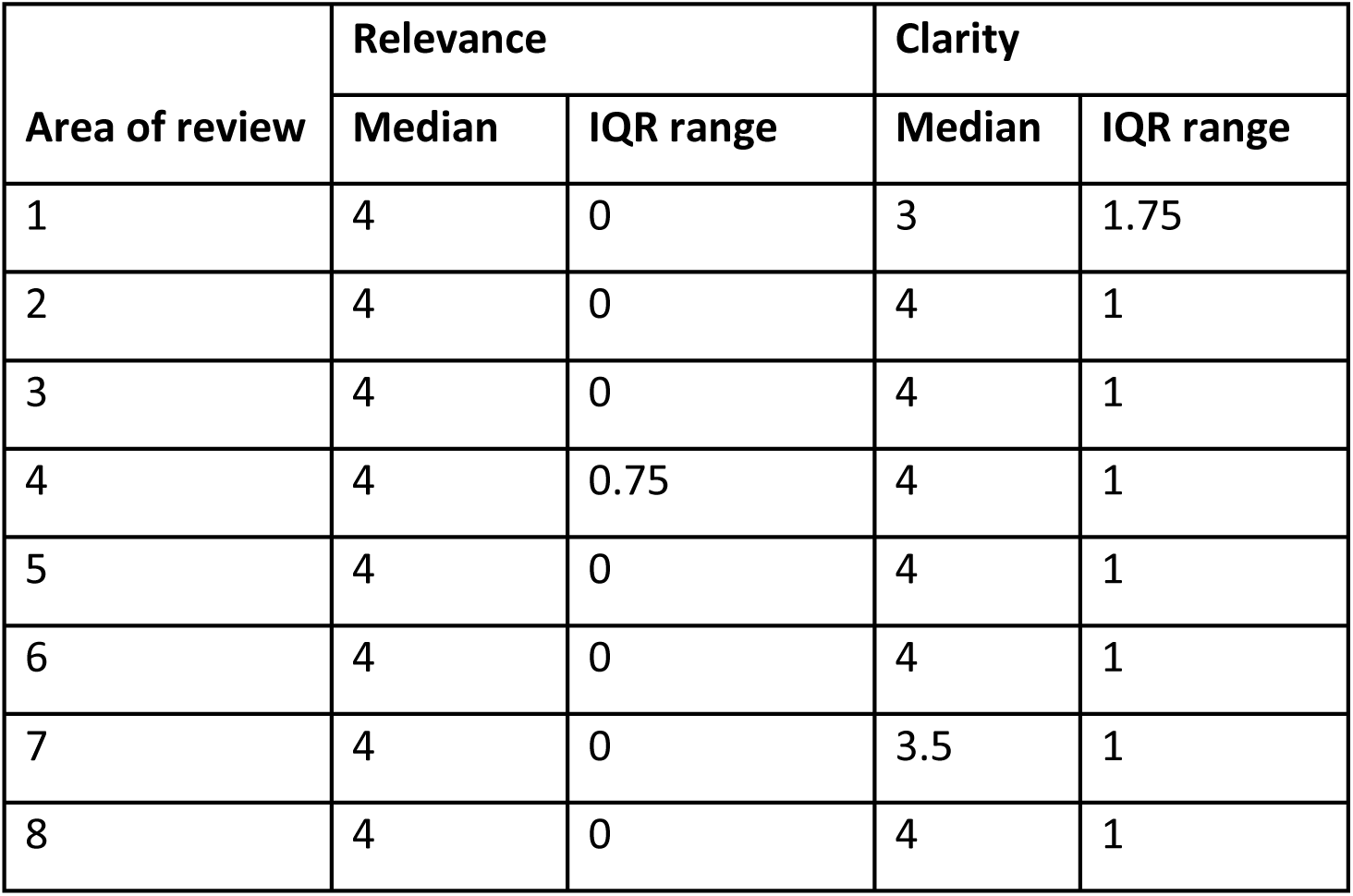
Round 1 results.

For example, area of review 1 was:

> “*People affected by incidents are meaningfully engaged and involved*”.

The description was:

> “*The report demonstrates evidence that all those affected by the incident such as staff, patients, families and carers have been actively listened to and emotionally supported where required (i.e. interviews and perspectives of those affected are included in the report*).”

This area of review was revised to:

> “*People affected by incidents are compassionately engaged and meaningfully involved*.”

The description was revised to:

> “*The report includes evidence that those affected by the incident such as staff, patients, families and carers have been actively listened to and supported (i.e. the perspectives of those affected, and how they have been supported, are included in the report.*)”

There were suggestions for the inclusion of additional areas of review. Following discussion within the team, none were taken forward because the points made were already included in the existing areas of review or that the points made were beyond the remit of the tool. For example, suggestions to include assessing whether appropriate people had been involved in the investigation and that the report demonstrates impartiality in the investigations. No areas of review were removed at the end of this round.

#### Round 2 Results

In round 2, participants were asked to only rate on the clarity of area of review 1, which did not reach consensus. At the end of round 2, consensus was reached on the clarity of area of review 1, with a median of 4 and IQR range of 1. Participants were given the opportunity to, and provided further comments on, the relevance and clarity of the wordings in the tool. The research team considered each comment and made minor changes to the wordings in 4 of the 8 areas of review. The final tool can be accessed and downloaded here: https://www.hssib.org.uk/education/learning-response-review-and-improvement-tool/

## Discussion

A healthcare safety investigation report tool was co-designed with ‘expert’ panel members. The content of the tool was validated with current users. The tool contains 8 areas of review, designed to support self-assessment and improvement of written safety investigations reports and learning reviews. The tool can also support organisational quality assurance or oversight mechanisms for monitoring, evaluating and improving the standard of reports. As written reports can act as a proxy measure for the quality of investigations, improving the standard of reports may inform the learning and associated action from patient safety investigations.

### Comparison with the existing literature

We are aware of previous studies reporting the development and use of a structured formative tool for reviewing and providing feedback on the quality of patient safety event reports. This research was conducted in the general medical practice setting in the west of Scotland and focused on development of a peer review system of written analyses of significant events submitted by family doctors as part of their continuing professional development obligations and also if they undertook educational supervisor training [26–27, 35–36]. Additionally, the World Health Organisation’s Concise Incident Analysis Tool [15] is previously reported to be applied by the Dutch Healthcare Inspectorate “to judge the quality of a sentinel event analysis reports” [10]. However, the question-set and scoring system differ significantly from our tool. For example, there is a reductionist focus on ‘root causes’ which has come in from recent significant global criticism [37–48], while NHS England’s PSIRF policy has moved on from that type of thinking and approach [33].

Overall, the tool aims to address many of the key challenges related to reviewing and learning from patient safety incident reports that are routinely reported across healthcare [43]. For example the direct and indirect use of blame and pejorative language, the lack of a ‘systems approach’ taken, and the weakness of recommendations for improvement that can be made [19–20].

### Study strengths and limitations

Key strengths of this study include the use of the modified-Delphi technique which is useful where published evidence to build consensus on a given topic is limited. The study involved a collaboration of multiple, informed and highly experienced participants from a range of regional and national healthcare organisations, academic bodies, and commercial enterprises with specialist knowledge of the topic area. While the consensus building process was judged to be rapid and robust using a recognised method and online tools for convenience, greater rigour might have been achieved through face-to-face discussions and contributions.

### Implications for policy, practice, education and research

The tool has multiple potential uses. For example,

1. To facilitate self-assessment and improvement by report writers
2. To provide formative and developmental feedback on reports by reviewers
3. To inform training designed to support systems thinking and reporting of and learning from healthcare safety investigations;
4. To provide senior leaders, executive teams and board members with valuable insight into the strengths and weaknesses of safety investigation reports, and by proxy, safety investigations.
5. To strengthen oversight and quality assurance processes related to healthcare safety investigations and reports.

Furthermore, while the focus of the tool is at the organisational safety investigation level with an initial focus on related written reports, it may also have potential utility in team-based learning responses to safety incidents. For example, the tool does not have to be applied to a written report but could be adapted and used as a simple ‘good practice’ guide or checklist by those in healthcare who chair or moderate multidisciplinary Mortality and Morbidity meetings and similar where patient safety incidents are routinely analysed (Box 1). It could also be used in routine team learning activities across all care sectors where patient safety and service quality issues are discussed, analysed and conclusions drawn.

**Box 1.**
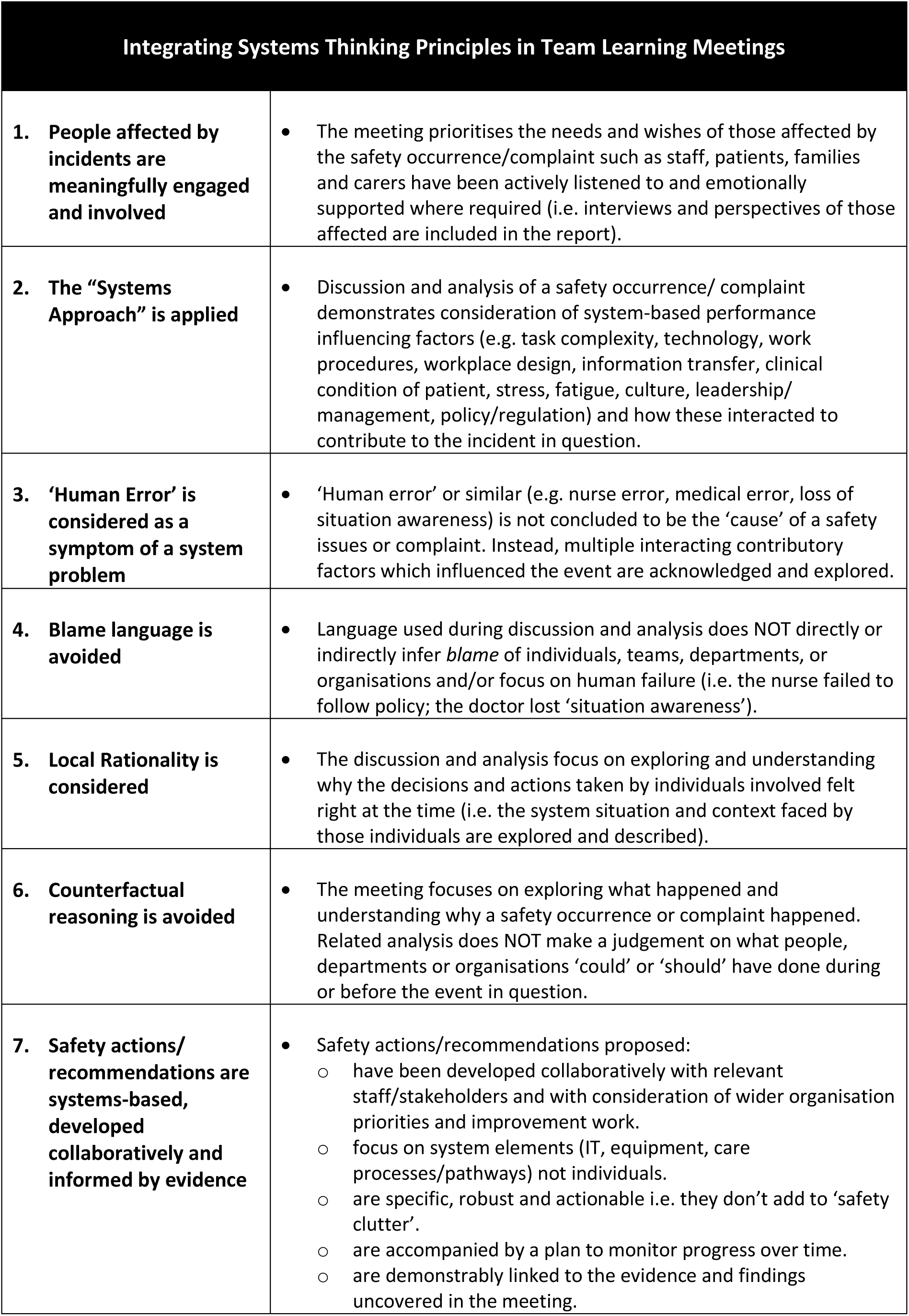
Guidance for Reviewing Patient Safety Incidents at Multidisciplinary Team Meetings.

Future work could include assessing the impact of using the tool on outcome measures such as the quality and variability of written reports. In addition, research could also be done to assess how the tool impacts healthcare safety investigation and report writing practice at the personal, team and organisational levels. Incorporating the tool with a large language model to automate the review process may make its use more efficient and offer a cost-benefit and is also worth future consideration.

Currently, the tool is included in a protocol for a multistage, mixed-methods study aimed at improving health system responses when patients are harmed [49]. In terms of its cross-industry potential, the tool has also been successfully applied to selected safety investigations undertaken in the international mining sector (Personal Communication, Nicola Steevenson). We encourage those with a professional interest in this area in the UK and internationally to use the tool in their own care sectors. The tool is freely available and can be accessed and downloaded here: <u>Link</u>

## Conclusion

We co-designed and validated the content of a safety investigation report writing tool that can be used in all healthcare settings. There are 8 areas of review that support a “systems-based” approach to healthcare safety investigations and learning. The tool has multiple uses ranging from self-assessment for report writers to facilitating oversight of the quality of healthcare safety investigation reports. Future research and evaluation work could focus on providing evidence of the tool’s overall utility, including potentially applying it to the formative review of high-profile national and international include safety investigation reports.

## Footnotes

### Contributors

PB, MO, TH and RL planned the study. PB, RL and MO conducted the study. All authors contributed to tool development and the critical review and writing of the manuscript. PB submitted the study.

## Patient consent

Not required.

## Ethics approval

Phase 1 of the study did not require ethical approval under the UK Research Ethics Governance Framework (2021). Phase 2 of the study received a favourable ethical opinion by the University of Reading School of Chemistry, Food and Nutritional Sciences and Pharmacy internal ethics review committee (study no: 16/2024 Amendment date 08/07/2024).

## Data Availability

All data produced in the present work are contained in the manuscript

